# Full genome viral sequences inform patterns of SARS-CoV-2 spread into and within Israel

**DOI:** 10.1101/2020.05.21.20104521

**Authors:** Danielle Miller, Michael A. Martin, Noam Harel, Talia Kustin, Omer Tirosh, Moran Meir, Nadav Sorek, Shiraz Gefen-Halevi, Sharon Amit, Olesya Vorontsov, Dana Wolf, Avi Peretz, Yonat Shemer-Avni, Diana Roif-Kaminsky, Na’ama Kopelman, Amit Huppert, Katia Koelle, Adi Stern

**Affiliations:** School of Molecular Cell Biology and Biotechnology, George S. Wise Faculty of Life Sciences, Tel Aviv University, Israel; Department of Biology, Emory University, Atlanta, GA, USA; Population Biology, Ecology, and Evolution Graduate Program, Laney Graduate School, Emory University, Atlanta, GA, USA; Microbiology laboratory, Assuta Ashdod University-Affiliated Hospital. Ashdod, Israel; Clinical Microbiology Laboratory, Sheba Medical Center, Ramat-Gan, Israel; Clinical Virology Unit, Hadassah Hebrew University Medical Center, Jerusalem, Israel; The Azrieli Faculty of Medicine, Bar-Ilan University, Safed, Israel; Clinical Microbiology Laboratory, The Baruch Padeh Medical Center, Poriya, Tiberias, Israel; Clinical Virology Laboratory, Soroka Medical Center and the Faculty of Health Sciences, Ben-Gurion University of the Negev, Beer-Sheva, Israel; Microbiology Division, Barzilai University Medical Center, Ashkelon, Israel; Department of Computer Science, Holon Institute of Technology, Holon, Israel; Bio-statistical and Bio-mathematical Unit, The Gertner Institute for Epidemiology and, Health Policy Research, Chaim Sheba Medical Center, Tel Hashomer, 52621 Israel; School of Public Health, the Sackler Faculty of Medicine, Tel-Aviv University, Tel Aviv, 69978 Israel; Edmond J. Safra Center for Bioinformatics at Tel Aviv University

## Abstract

Full genome sequences are increasingly used to track the geographic spread and transmission dynamics of viral pathogens. Here, with a focus on Israel, we sequenced 212 SARS-CoV-2 sequences and use them to perform a comprehensive analysis to trace the origins and spread of the virus. A phylogenetic analysis including thousands of globally sampled sequences allowed us to infer multiple independent introductions into Israel, followed by local transmission. Returning travelers from the U.S. contributed dramatically more to viral spread relative to their proportion in incoming infected travelers. Using phylodynamic analysis, we estimated that the basic reproduction number of the virus was initially around ~2.0-2.6, dropping by two-thirds following the implementation of social distancing measures. A comparison between reported and model-estimated case numbers indicated high levels of transmission heterogeneity in SARS-CoV-2 spread, with between 1-10% of infected individuals resulting in 80% of secondary infections. Overall, our findings underscore the ability of this virus to efficiently transmit between and within countries, as well as demonstrate the effectiveness of social distancing measures for reducing its spread.

## INTRODUCTION

In December 2019 an outbreak of severe respiratory disease was identified in Wuhan, China (Huang, et al. 2020). Shortly later, the etiological agent of the disease was identified as severe acute respiratory syndrome coronavirus 2 (SARS-CoV-2) (Zhou, et al. 2020; Zhu, et al. 2020), and the disease caused by the virus was named coronavirus disease 19 (COVID-19). The virus has since spread rapidly across the globe, causing a WHO-declared pandemic with social and economic devastation in many regions of the world. The infectious disease research community has quickly stepped up to the task of characterizing the virus and its replication dynamics, describing its pathogenesis, and tracking its movement through the human population. Parameterized epidemiological models have been particularly informative of how this virus has spread with and without control measures in place (e.g., Tian, et al. 2020), and have been used to project viral spread both in the short-term (Flaxman, et al. 2020) and in the more distant future (Kissler, et al. 2020).

Along with epidemiological analysis based on case reports and COVID-19 death data, sequencing of viral genomes has become a powerful tool in understanding and tracking the dynamics of infections (Volz, et al. 2013; Gardy and Loman 2018). So-called “genomic epidemiology” allows for effective reconstruction of viral geographical spread as well as estimation of key epidemiological quantities such as the basic reproduction number of a virus, its growth rate and doubling time, and patterns of disease incidence and prevalence. Such insights have been used to inform policy makers during various pathogen outbreaks, as occurred for example in the 2014-2016 outbreak of Ebola virus in West Africa (Khoury, et al. 2018; Armstrong, et al. 2019), and during this current SARS-CoV-2 pandemic (Bedford, et al. 2020; Fauver, et al. 2020).

Here, we set out to sequence SARS-CoV-2 from samples across the state of Israel, with the aim of gaining a better understanding of introductions of the virus into Israel, spread of the virus inside the country, and the epidemiology of the disease, including (a) the basic reproduction number of the virus before and after social distancing measures were implemented, and (b) the extent of viral superspreading within Israel. We sought to gain this understanding within the context of existing epidemiological knowledge, including that the first confirmed cases of SARS-CoV-2 infection in Israel were reported in early-February, followed by many identified SARS-CoV-2 cases in travelers returning to Israel mainly from Europe and the United States. Growth in the number of verified cases rapidly ensued, which led to increased measures of social distancing, including the cessation of passenger flights to Israel, school closure, and eventually a near complete lockdown across the entire state of Israel. Quarantining of returning travelers from Europe was implemented between February 26 and March 4, 2020, and subsequently all incoming travel to Israel was arrested on March 9. In the meantime, the rate of testing was ramped up, eventually reaching a rate of more than 1,500 tests per million people per day. The reported daily incidence and reported numbers of daily severe cases peaked around mid-April and have dropped steadily since then. Despite this knowledge, many questions remain: Which of the multiple SARS-CoV-2 introductions resulted in sustained local transmission? How did the virus spread across the state? What was the magnitude of the virus’s reproduction potential within Israel, and to what extent did control measures mitigate its spread? Here, through a comprehensive set of phylogenetic and phylodynamic analyses, we quantitatively address these questions.

## RESULTS & DISCUSSION

In order to gain a better understanding of the dynamics of SARS-CoV-2 spread into and within Israel, we sequenced the virus from a cohort of patients representing a random sample across Israel, resulting in 212 full-genome SARS-CoV-2 sequences (Methods). A total of 224 unique single nucleotide variants (SNVs) were identified between the Wuhan reference sequence and this set of sequences from Israel. Figure 1 shows the distribution of identified SNVs along the genome and their counts in the sequenced samples. Of these SNVs, 141 were non-synonymous, 72 were synonymous, and the remaining 11 were in non-coding regions. One of the most abundantly detected SNVs was a non-synonymous variant D614G found in the spike protein, which was present in 90% of the sequences. This variant has generated much interest as it has been reported to potentially increase the transmissibility of the virus (Korber, et al. 2020). Of note however, the alternative hypothesis that the observed increases in this variant’s frequency is due to demographic considerations and genetic drift has not been ruled out.

**Figure 1.**
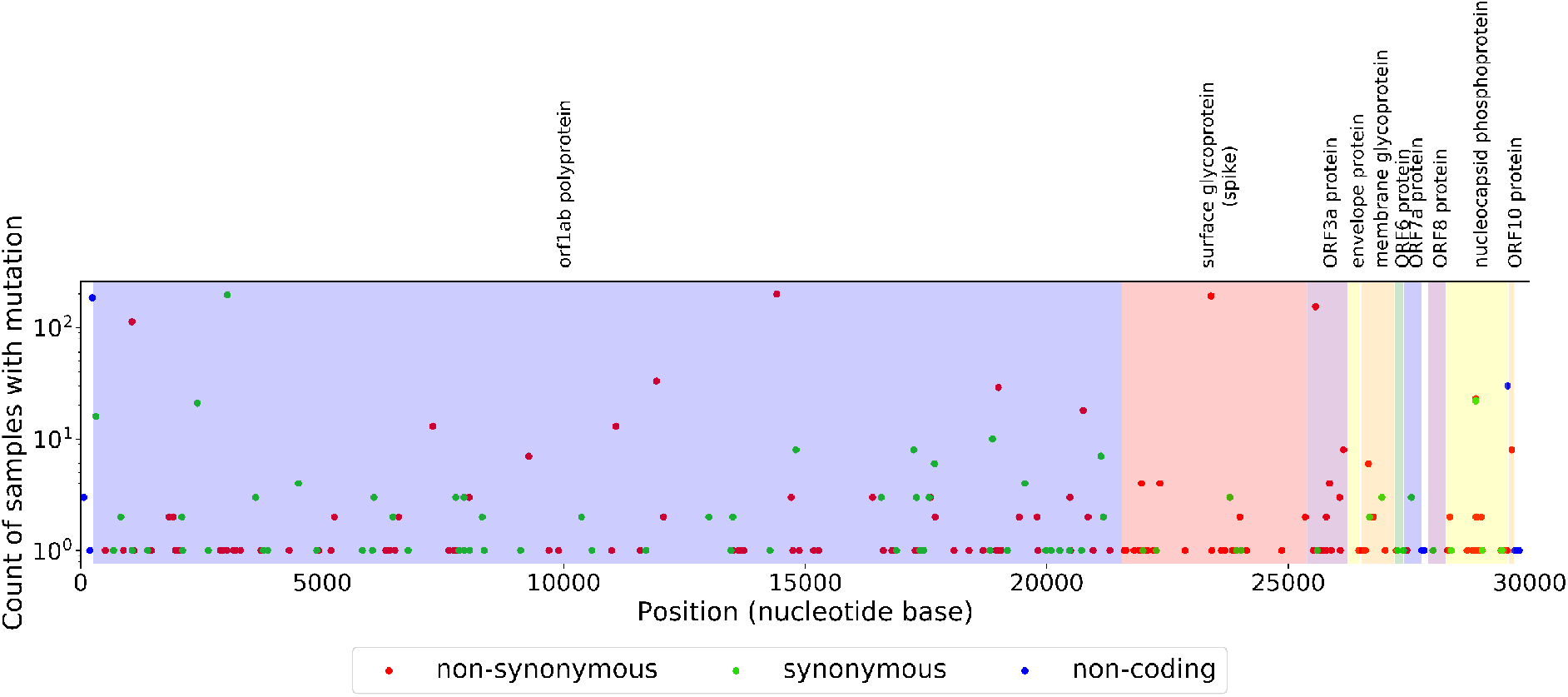
Variation found in sequenced samples from Israel. Counts of identified SNVs across the SARS-CoV-2 genome.

We also found five different high confidence genomic deletions, spanning between one and eighteen nucleotides (Fig. 2) (Methods), each of which was found in one to two samples. Three of these five deletions occurred in multiples of three and were in-frame deletions or affected non-coding regions. Of the remaining two deletions, deletion #3 spanned ten nucleotides, and likely prevents the translation of ORF7a. Deletion #4 occurred at the end of ORF8 and causes the replacement of the last amino acid with an additional five amino acids. Notably, an 81 nt in-frame deletion in ORF7a has been previously reported (Holland, et al. 2020), as has been a 382 nt deletion in ORF8 (Su, et al. 2020), suggesting that the virus is to some extent tolerant to deletions in these ORFs.

**Figure 2.**
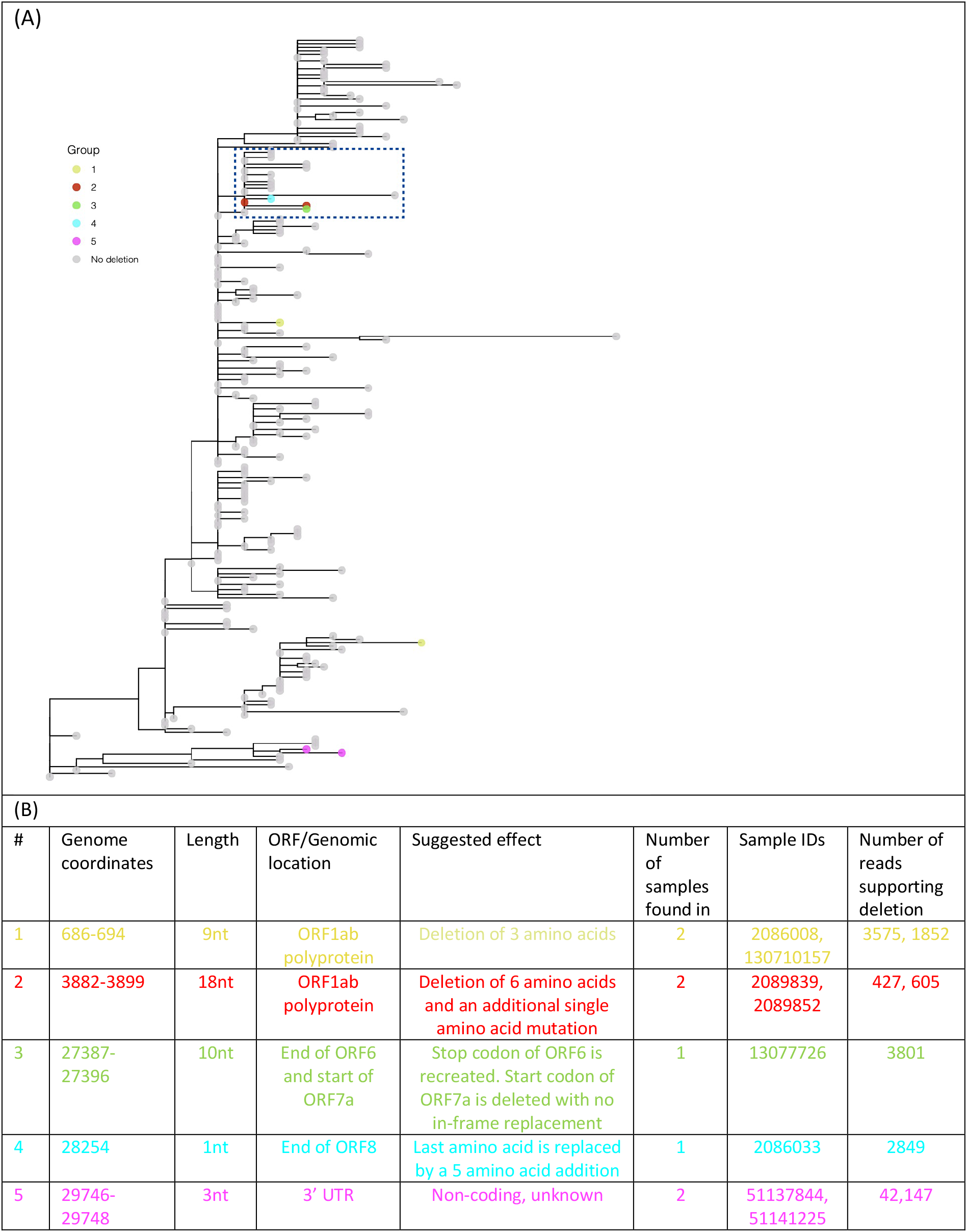
Deletions found in Israeli samples. (A) Maximum-likelihood tree of Israeli sequences highlighting sequences found with deletions that are color-coded and described in (B). A clade with three independent deletions occurring in four samples is boxed. The phylogeny displayed was created with the ggtree package (Yu, et al. 2017).

When focusing on deletions that occurred in two samples, we noted that the two samples bearing deletion #1 were located in very remote clades on the phylogeny, suggesting that the deletion occurred two independent times. Deletion #5 on the other hand was present in two related samples that were sampled five days apart from each other. Deletions #2, #3 and #4 revealed an intriguing pattern: three independent deletions (one of which was present in two samples) were all part of the same clade that included eighteen samples (Fig. 2). One non-synonymous SNV defined this clade: S2430R in ORF1b which affects the non-structural protein NSP16. This protein has been reported to be a 2’O-methyl-transferase that enhances evasion of the innate immune system (Menachery, et al. 2014).

While further in-depth investigation of SARS-CoV-2 indels is clearly needed, at this point we conjecture that the deletions we detected are neutral or to some extent deleterious, and that deletions in SARS-CoV-2 are likely to occur frequently given the number of deletions detected in our samples.

### Origins and transmission patterns in Israel

We next set out to explore patterns of SARS-CoV-2 introduction into Israel. Figure 3 shows the time-resolved phylogeny inferred using the 214 Israeli sequences (212 sequenced here and two additional ones sequenced previously) in addition to 4,693 representative sequences from across the world. This phylogeny allowed us to characterize the major viral clades circulating within Israel and to infer the geographic sources and timing of virus introductions into the state. We found multiple introductions into Israel from both the U.S and Europe, the latter including mainly the U.K., France and Belgium. Over 70% of the clade introductions into Israel (confidence intervals ranging from ~50% to ~80%, see Methods) were inferred to have occurred from the U.S., while the remaining were mainly from Europe. During the entire epidemic in Israel, very close monitoring of all incoming infected travelers was imposed, and reports show that only ~27% of infected returning travelers were from the U.S. The discrepancy between these estimated proportions suggests that the travelers returning from the U.S. contributed substantially more to the spread of the virus in Israel than would be proportionally expected. This may have occurred due to the gap in policy that allowed returning non-European travelers to avoid quarantine until March 9, or due to different contact patterns of those who returned from the U.S. Moreover, this suggests that had flights from the U.S. been arrested at the same time that flights from Europe were arrested (between February 26 and March 4, instead of by March 9), a substantial fraction of the transmission chains in Israel would have been prevented.

**Figure 3.**
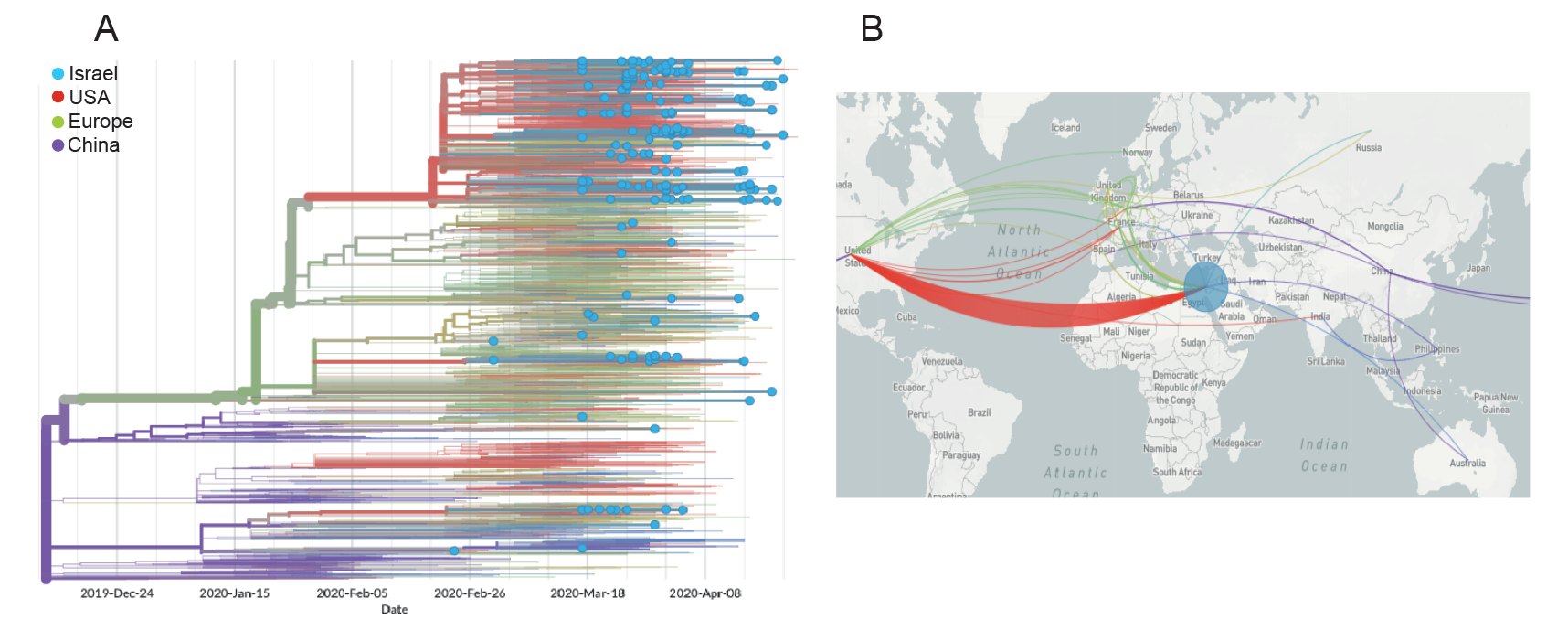
Patterns of SARS-CoV-2 introduction into Israel. (A) Time-resolved phylogeny inferred using viral sequences from Israel (blue tips) and around the world (tips without dots). Lineages are colored by inferred region of circulation. Phylogeographic analysis reveals multiple introductions into Israel, mainly from the U.S. (B) Map of phylogenetically inferred introductions into Israel highlighting the dominance of the U.S. and to a lesser extent Europe as the geographic sources of SARS-CoV-2 introductions into Israel. Figures generated using NextStrain (Hadfield, et al. 2018).

As the pandemic spread, entry into Israel was restricted, and local transmissions became dominant. Transmission patterns into and between six various geographical regions in Israel (North district, Tel Aviv district, South coast district, Jerusalem district, and South district) are shown in Figure 4. While most local transmission occurred inside defined regions, transmission among distinct regions was also observed, such as, for example, movement between Jerusalem and the north district of Israel.

**Figure 4.**
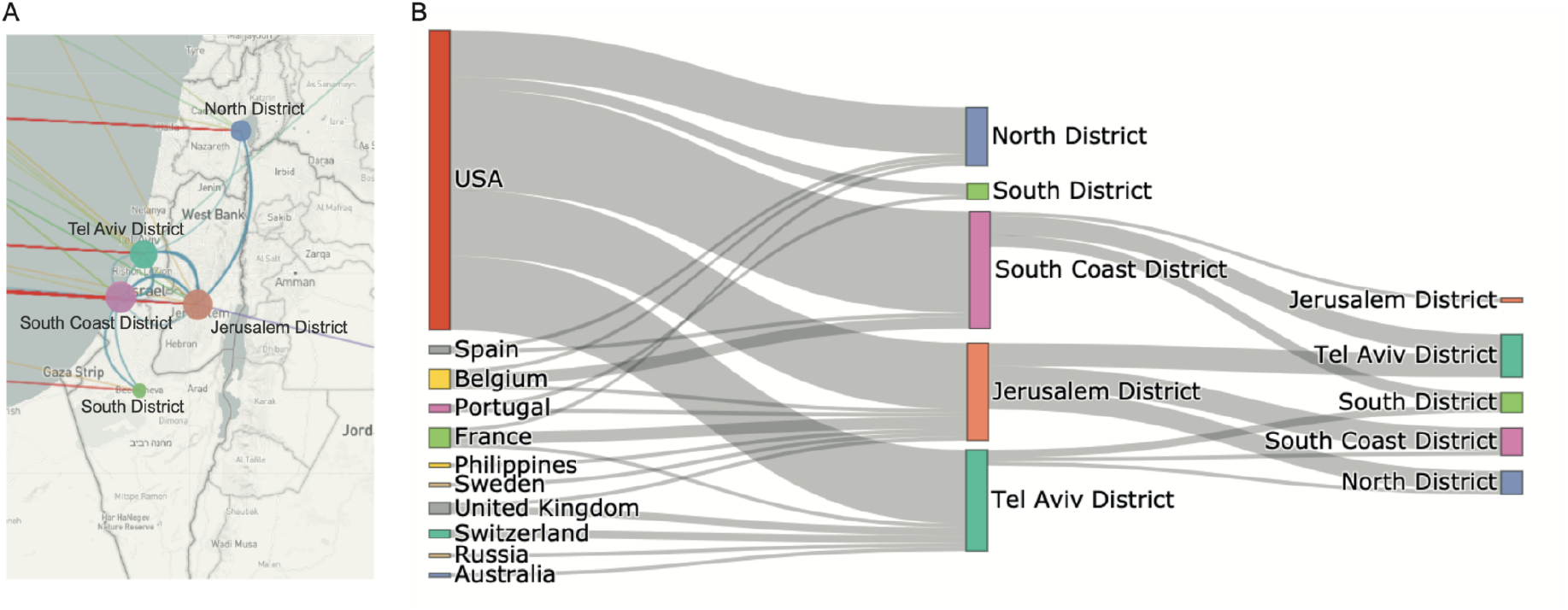
Spread of SARS-CoV-2 into and within Israel. (A) Map of Israel with geographic locations of samples, and inferred spread into Israel (colored lines) and inside Israel (blue lines). (B) Alternative view of spread into and inside Israel. Each line represents a transmission event inferred based on the phylogeny. Thicker lines indicate multiple transmission events.

### Phylodynamic modeling of viral spread in Israel

To estimate the basic reproduction number of SARS-CoV-2 in Israel initially and then following the implementation of social distancing measures, we performed coalescent-based phylodynamic inference using PhyDyn (Methods). We note that existing phylodynamic analyses of SARS-CoV-2 have shown that the effective reproduction number *R_E_* of the virus has decreased over time, as quarantine and social distancing measures have been implemented within specific regions (Danesh, et al. 2020; Vaughan, et al. 2020; Volz, Baguelin, et al. 2020). However, many of these analyses have to date modeled reductions in *R_E_* as stemming from the depletion of susceptible individuals (Volz, Baguelin, et al. 2020), rather than from reductions in *R*_0_, the latter of which would be consistent with lowering of contact rates. Other analyses, particularly those that use the birth-death model approach for phylodynamic inference, have allowed for changes in *R*_0_ over time (Danesh, et al. 2020; Vaughan, et al. 2020), but cannot as easily accommodate structure in the infected host population (e.g., that some individuals are exposed but not yet infectious, and that transmission heterogeneity exists between infected individuals). Our phylodynamic analysis here, based heavily on existing coalescent-based model structures that have been applied to SARS-CoV-2 (Volz 2020), instead allows for this structure to be accommodated and for *R*_0_ to change in a piecewise fashion over time.

Our phylodynamic analysis assumes an underlying susceptible-exposed-infected-recovered (SEIR) epidemiological model for SARS-CoV-2 transmission dynamics and explicitly incorporates transmission heterogeneity (Methods). Recent epidemiological analyses have estimated considerable levels of SARS-CoV-2 transmission heterogeneity, with ~7-10% of infected individuals estimated to be responsible for 80% of secondary infections (Bi, et al. 2020; Endo, et al. 2020). Instead of assuming a given level of transmission heterogeneity for Israel, we instead performed phylodynamic inference of the SEIR model across a range of transmission heterogeneities ranging from *p_h_* = 1% to 20% of infected individuals being responsible for 80% of secondary infections. We estimated *R*_0_ prior to March 19 to be approximately 2.0 across this superspreading range, with estimates increasing towards *R*_0_ = 2.4 to 2.6 at extremely high levels of superspreading (*p*_h_ = 1-2%) (Figure 5A). Across this superspreading range, we robustly estimated that quarantine measures had the effect of reducing *R*_0_ by approximately two-thirds (α = ~30%; Figure 5B; see Table S2 for model parameter priors and estimated values).

**Figure 5.**
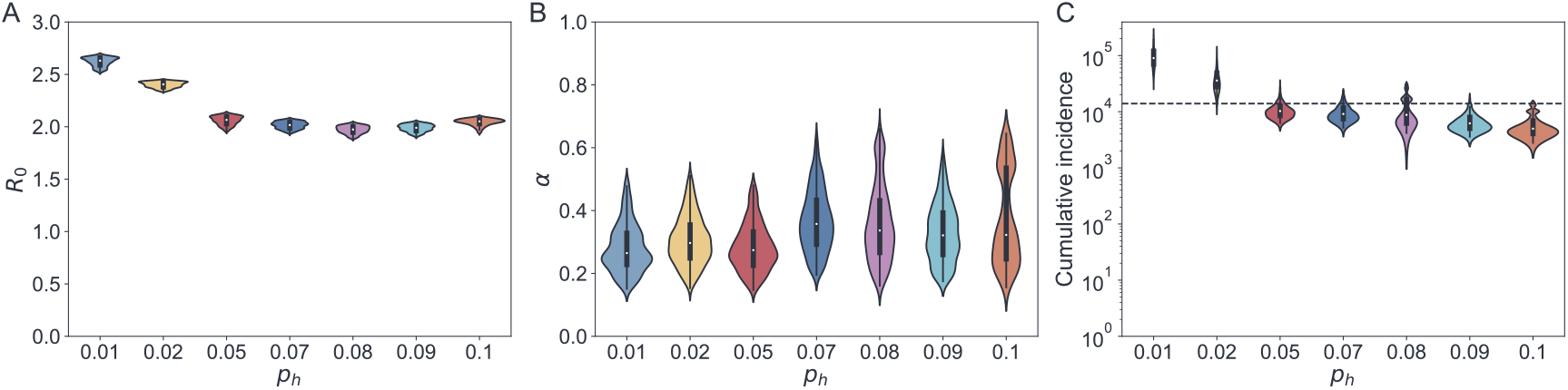
Estimated epidemiological parameters across different levels of transmission heterogeneity. The parameter *p_h_* gives the fraction of infected individuals that are responsible for *P =* 80% of secondary infections. Higher *p_h_* values correspond to less transmission heterogeneity. (A) Estimated *R*_0_ in Israel prior to March 19, 2020. (B) Estimated fraction by which *R*_0_ in Israel changed after March 19. (C) Estimated cumulative number of infected individuals in Israel on the date of the last sampled sequence (April 22, 2020). An infected individual is assumed to contribute to cumulative incidence at the end of their infectious period. Horizontal dotted line at *N* = 13,942 shows the cumulative number of reported cases on April 22, 2020 as given by the ECDC (https://opendata.ecdc.europa.eu/covid19/casedistribution/csv). In (A)-(C), only values that fall within the 95% highest posterior density intervals are shown.

Figure 5C shows the cumulative number of SARS-CoV-2 cases by April 22, estimated by our phylodynamic analyses across the considered superspreading range. Estimates of the cumulative number of cases is highly sensitive to the level of assumed transmission heterogeneity, particularly at high levels of superspreading (*p_h_* = 1-5%). Comparison between these inferred cumulative cases and reported case numbers (dotted line in Figure 5C) indicates that SARS-CoV-2 transmission dynamics were driven by an extremely high level of viral superspreading. Specifically, if we assume almost complete case reporting, our phylodynamic analysis indicates that between 5-9% of infections are responsible for 80% of secondary infections; with lower assumed levels of case reporting, between 1-5% of infections would be responsible for this 80% of secondary infections.

Phylodynamic analysis further allows us to visualize inferred epidemiological dynamics. In Figure 6, we show inferred patterns of prevalence (Fig. 6A) and incidence (Fig. 6B) for three different assumed levels of viral superspreading. Inferred patterns of prevalence corroborate epidemiological findings that the number of cases started to decline in early April. Inferred patterns of cumulative incidence indicate that reporting rates were initially low, but improved considerably over the time course of viral spread. The ‘leveling off of cumulative incidence around late March/early April observed in both the reported case data and in our inferred epidemiological dynamics, ground-truthing the results of our phylodynamic analyses.

**Figure 6.**
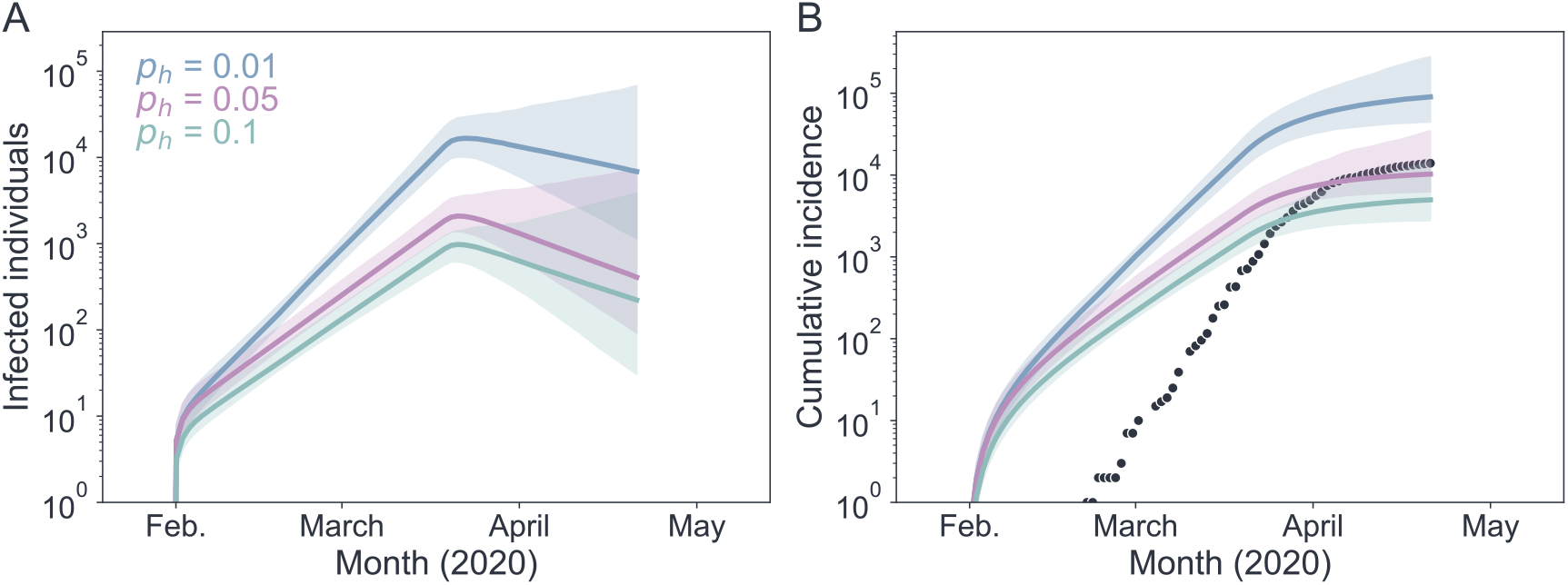
Epidemiological dynamics inferred using phylodynamic analysis. (A) Estimated number of currently infected individuals (*I*_l_ + *I*_h_) over time. (B) Estimated cumulative number of infected individuals. An infected individual is assumed to contribute to cumulative incidence at the end of their infectious period. Black dots show the cumulative number of reported cases in Israel over time, as given by the ECDC. Model-predicted numbers of infected individuals include both classes of infected individuals (*I*_l_ + *I*_h_). In (A) and (B), lines show median estimates of models with different levels of transmission heterogeneity: *p_h_ =* 0.01 (blue), 0.05 (pink), and 0.10 (green). Shaded regions show the 95% highest posterior density.

### Conclusions

Overall, our findings highlight the use of genomic data to effectively track the spread of an emerging virus using phylogenetic and phylodynamic approaches that have been developed to study viral outbreaks. We have hereby succeeded in tracking the main transmission chains that led to SARS-CoV-2 spread in Israel, and applied phylodynamic analysis to infer key epidemiological parameters governing its spread. Our results suggest that superspreading events are a main feature of SARS-CoV-2 spread, suggesting that focused measures to reduce contacts of select individuals/social events could dramatically mitigate viral spread. Finally our results highlight how global connectivity allows for massive introductions of a virus, and emphasize how border control and shelter-in-place restrictions are crucial for halting viral spread.

## METHODS

### Ethics statement

An exemption from institutional review board approval was determined by the Israeli Ministry of Health as part of an active epidemiological investigation, based on use of anonymous data only and no medical intervention. The study was further approved by the Tel-Aviv University ethics committee (approval 0001274-1).

### Details of samples & virus genome sequencing

With the aim of generating a random sample of viral infections across the entire country, a total of 213 samples were retrieved from six major hospitals in Israel spanning the entire geography of Israel from south to north (Table 1, Table S1).

**Table 1.**
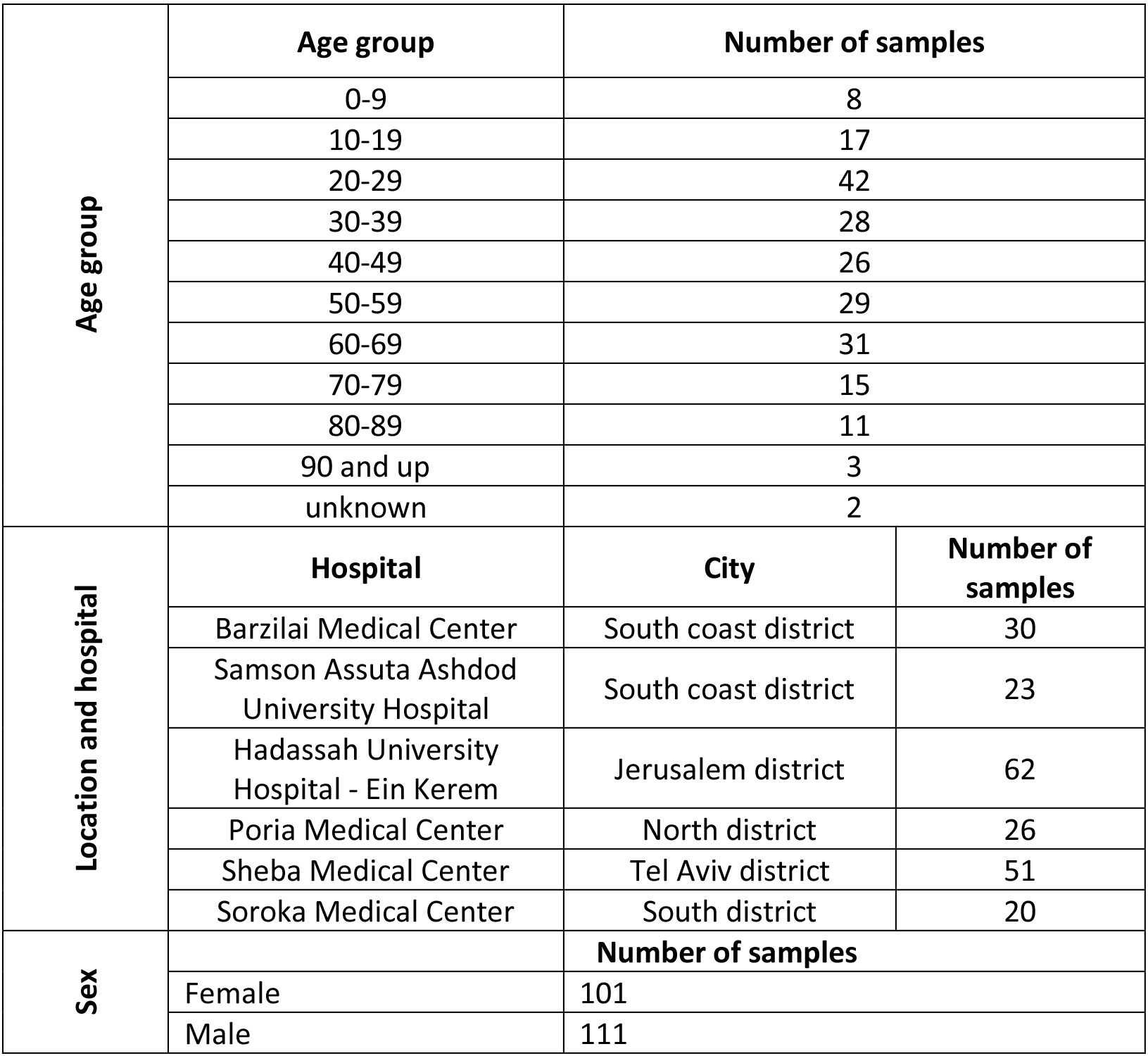
General statistics on samples collected and sequenced.

We obtained RNA extracted from nasopharyngeal samples. Sequencing was performed based on the V3 Arctic protocol (https://artic.network/ncov-2019). Briefly, reverse transcription and multiplex PCR of 109 amplicons was performed, and adapters were ligated to allow for sequencing. All samples were run on an Illumina Miseq using 250-cycle V2 kits in the Technion Genome Center (Israel).

### Determining genome consensus sequences

Sequencing reads were trimmed using pTrimmer, a multiplexing primer trimming tool (Zhang, et al. 2019), and then aligned to the reference genome of the SARS-CoV-2 (GenBank ID MN908947) using our AccuNGS pipeline (Gelbart, et al. 2019), which is based on BLAST (Altschul, et al. 1997), using an e-value of 10^-9^. The pipeline allows for consensus determination and variant calling. We considered substitutions at the consensus sequence (as compared to the reference) only if a given base was present in 80% of the aligned reads, and five or more reads aligned to the reference; bases where the majority of reads showed a substitution but that did not fulfill these two conditions were deemed uncertain. Similarly positions to which no reads were mapped were also deemed uncertain, and such positions were assigned with an “N”. All deletions were manually verified: (a) over 98% of the reads covering the deletion site mapped to both ends of the deletion (i.e, bore evidence of the deletions), (b) the deletion was based on over 40 independent reads (on average >1,000 reads), and (c) coverage was high at both ends of the deleted region. Only sequences that spanned 90% of the reference genome were retained, leading to the removal of one sequence (Table S1), and hence a new set of 212 Israeli sequences was generated here. Another two Israeli sequences already available on GISAID were added to the phylogenetic analysis, leading to a total of 214 sequences from Israel.

The collection dates of the 214 Israeli sequences used in our analysis ranged from February 23 through April 22, 2020. The number of sequences is thus approximately 1.5% of the total number of reported cases on April 22.

### Phylogenetic analyses

All available full-length SARS-CoV-2 genomes from outside of Israel (a total of 16,403 sequences) were retrieved from GISAID on May 5, 2020. All sequences from a non-human host as well as sequences with incomplete sampling date (YYYY-MM or YYYY-MM-XX) or a high level of uncertainty (>10% ambiguous bases marked as N) were removed. All available sequences were then down-sampled to 4,693 representative sequences across the globe using the latest build of NextStrain ncov pipeline (Hadfield, et al. 2018; Hadfield, et al. 2019) (https://github.com/nextstrain/ncov). 1195 of these 4,693 sequences were from the U.S., while 1991 were from Europe. The 212 new Israeli sequences were added to the tree.

### Confidence in numbers and fractions of importation events

Confidence in the relative number of importation events from the U.S. vs. Europe was assessed using two measures of confidence intervals, which were aimed at testing whether the set of exogenous sequences was biased, or whether the set of Israeli sequences was biased. First, we generated 1,000 samples of the exogenous (non-Israeli) sequences using a bootstrap approach and assessed the fraction of importation events in each sample. Second, we similarly bootstrapped the local (Israeli) sequences only and assessed the fraction of importation events in each sample. The reported confidence interval includes the lower bound and higher bound of both bootstrapping schemes.

### Down-sampling of global tree for phylodynamic analysis

Following the initial sampling of the global tree described above, we applied a second sampling specifically for the phylodynamic analysis. The down-sampling followed the recommended guidelines described previously for SARS-CoV-2 (Volz, Boyd, et al. 2020). We used two sampling techniques:

i. *Random time stratified sampling* – we sampled a total of 100 sequences from outside of Israel across *ν*=5 time intervals such that each time interval contained approximately 20 sequences.
ii. *Closest sequence match* – we will define *S_ISR_* as the set of all sequences from Israel. We sample the exogenous set of sequences from the global tree with the minimal cophenetic distance between each Israeli sequence belonging to *S_ISR_* as based on the maximum likelihood phylogeny. This allow including sequences closely related to sequences from Israel.

We next manually curated the sequences from Israel to ensure they represent a random sample across Israel. To this end we removed samples suspected to be from the same household, samples with consecutive identifiers, or identical samples with similar identifiers and similar dates. Only one sample from a given household was chosen randomly. This led to a removal of 6 sequences.

Following down-sampling and manual curation, a phylogenetic tree was inferred using the NextStrain pipeline (Hadfield, et al. 2018). The tree topology was validated as a legitimate representative of the global tree by performing 1,000 random samples containing 373 sequences from the global tree. The Kendall-Colijn metric (Kendall and Colijn 2016) was used to assess distance between each random sample and the original tree, allowing us to create a null distribution. The *λ* parameter, which determines the trade-off between topology and branch length, was set to zero, thus accounting for the tree topology alone. The significance of down-sampling procedure was thus obtained by comparing the Kendall-Colijn metric of the down-sampled tree to the null distribution.

### Rate of importations

As described previously, strong global connectivity can result in a high number of independent seeding events (Chinazzi, et al. 2020), and we thus aimed at generating the distribution of importation dates (Volz, Boyd, et al. 2020). This was achieved based on the time-resolved phylogeny, which was then used to estimate the date of initial importation as well as the date on which the rate of new importations dropped. Given a down sampled ML tree *T^ML^*, we re-estimate a forest of trees using IQ Tree (Nguyen, et al. 2015) 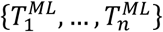 such that each tree 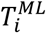 is generated by randomly resolving polytomies in *T^ML^*. Each 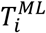 is used to produce a time-based tree, 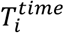 using TreeTime (Sagulenko, et al. 2018) with molecular clock rate *r~U*[0.0009 − 0.0015] substitutions/site/year. We deduce the distribution of all seeding events by taking the mid-branch date for each node leading to an Israeli tip for all trees. Both IQ Tree and TreeTime were executed using the Augur python package.

### Phylodynamic Analysis

Phylodynamic analyses were conducted using BEAST2 v2.6.2 (Bouckaert, et al. 2019) and PhyDyn v1.3.6 (Volz and Siveroni 2018). An HKY substitution model with a lognormal prior for *κ* with mean log(*κ*) = 1.0 and standard deviation of log(*κ*) = 1.25 was used. We assumed no sites to be invariant and used an exponential prior for *γ* with a mean of 1.0. A strict molecular clock with a uniform prior between 0.0007 and 0.002 substitutions/site/year was used. A uniform prior was used for nucleotide frequencies. The down-sampled maximum likelihood tree generated using IQ Tree was used as a starting tree.

PhyDyn is a coalescent-based inference approach implemented in BEAST2, allowing for the integration over phylogenetic uncertainty (Volz 2012; Volz and Siveroni 2018). The program requires specification of an underlying epidemiological model, as well as any priors on parameters that will be estimated. In line with recent analyses (sarscov2.phylodynamics.org), we assumed that the epidemiological dynamics of SARS-CoV-2 were governed by Susceptible-Exposed-Infected-Recovered *(SEIR)* dynamics. Transmission heterogeneity has previously been described for viral pathogens including SARS-CoV-1 (Lloyd-Smith, et al. 2005) and appears to be important in the transmission dynamics of SARS-CoV-2 (Bi, et al. 2020; Endo, et al. 2020). To account for the possibility of transmission heterogeneity, as in previous work (sarscov2.phylodynamics.org), we modeled two classes of infected individuals: one with low transmissibility *I*_I_ and one with high transmissibility *I*_h_. Mathematically, the epidemiological model is given by

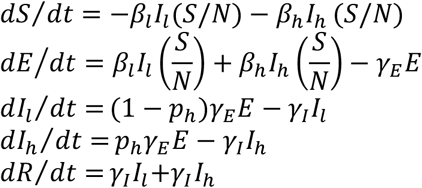

We set as fixed the host population size to the population size of Israel, according to the European Centre for Disease Prevention and Control (*N* = 8,883,800), the average duration of time an individual spends in the exposed class (1/*γ_E_* = 3 days), and the average duration of time an individual spends in the infected (infectious) class (1/*γ_I_* = 5.5 days). These rates are based on a study that inferred transmissibility over the course of infection based on data from established SARS-CoV-2 transmission pairs (He, et al. 2020). *R*_0_ in this model is given by *(β_h_p_h_. + β_l_*(1−*p_h_))/γ_I_*, where *p_h_* is the fraction of exposed individuals who transition to the *I_h_* class. Instead of independently parameterizing *β_h_* and *β_l_*, we defined (as in previous work) the relative transmissibility of infected individuals in the *I_h_* and *I*_I_ classes by the parameter *τ = β_h_/β_l_*. We defined a parameter *P* as the fraction of secondary infections that were caused by a fraction *p_h_* of the most transmissible infected individuals and set *P* to 0.8. Based on set values of *P* and *p_h_*, we calculated *r* as 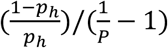. As such, we could easily parameterize the model across various levels of transmission heterogeneity, with a fraction *p_h_* of infected individuals being responsible for 80% of secondary infections. Existing epidemiological analysis indicate that *p_h_* is on the order of 7-10% (Bi, et al. 2020; Endo, et al. 2020) indicative of even more transmission heterogeneity than given by the 20/80 rule (Woolhouse et al., PNAS, 1997). We focused on the range of *p_h_* between 1-10% in our phylodynamic analyses.

Again based on existing analyses (sarscov2.phylodynamics.org), we included an external reservoir, *Y*, in our analysis to allow for multiple introduced clades into Israel to be jointly considered. We assume symmetric migration of individuals in the *E, I_I_*, and *I_h_* classes in and out of Israel based on a per capita migration rate *η*. An exponential prior with a mean of 10 and an upper limit of 10 was used for *η*. Based on the results of the importation analysis described above, we assumed that the migration rate decreased to 25% of its original value after March 20, 2020. As migration is assumed to be symmetrical in and out of Israel it does not affect the focal *SEIR* model dynamics, however, it influences the probability that a given lineage’s geographic state is assigned to Israel. We fix the rate of removal of individuals from *Y* to be 1/(1/*γ_E_ +* 1/*γ_I_)* and estimate the rate of entry into *Y, ρ*, using a lognormal distribution with mean log(*ρ*) = 3.6 and standard deviation log(*ρ*) = 1.

In our model, we estimated a piecewise *R*_0_ by estimating an initial *R*_0_ that was in effect until March 19, 2020 slightly after strong social distancing measures were implemented, along with a factor *α* by which *R*_0_ changed on March 19. We set the prior on *R*_0_ to a lognormal distribution with mean log (*R*_0_) of 1.5 and standard deviation of log (*R*_0_) of 0.5. We set the prior on *α* to be uniform between 0 and 2, thereby allowing *R*_0_ to either increase, decrease, or remain unchanged after March 19. An exponential prior with mean 1.0 was used for the initial size of the *E* class. The *I_I_* and *I_h_* were assumed to be negligibly small at the beginning of the *SEIR* dynamics. The PhyDyn *t*_0_ parameter was set to 2019.7 and a constant population size coalescent model was used prior to this date when proposed trees had earlier root dates. *SEIR* dynamics were assumed to begin on February 1^st^. Sequences sampled from Israel were randomly assigned to *I_h_* with probability *p_h_* and to *I_I_* with probability 1 − *p_h_*. XML files to run both BEAST2 and PhyDyn were generated using a custom Python 3 script which was designed to edit a template XML file originally generated with BEAUti and manually edited. MCMC chains for each parameter set were run for at least 1.9 million steps. Convergence was assessed based on visual inspection of parameter traces. Longer MCMC chains and additional replicates are currently being conducted. The first 50% of MCMC steps were discarded as burn-in. BEAST2 log and tree files were combined using LogCombiner. Maximum clade credibility trees were generated using TreeAnnotator. BEAST2 and PhyDyn outputs were visualized using Python 3, Matplotlib (Hunter 2007), Seaborn, and Baltic (https://github.com/evogytis/baltic).

## Data Availability

The assembled SARS-CoV-2 genomes (consensus sequences) were uploaded to GISAID (Table S1). Submission of the raw sequencing data to Sequence Read Archive (SRA) is pending. Code for phylodynamic analysis and model XML configuration, as well as scripts to analyze outputs are available on github.

https://github.com/SternLabTAU/SARSCOV2NGS

## Data and code availability

The assembled SARS-CoV-2 genomes (consensus sequences) were uploaded to GISAID (Table S1). Submission of the raw sequencing data to Sequence Read Archive (SRA) is pending. Code for phylodynamic analysis and model XML configuration, as well as scripts to analyze outputs are available at: https://github.com/SternLabTAU/SARSCOV2NGS.

## Acknowledgements

We wish to thank Dr. Boaz Lev at the Israeli Ministry of Health and Dr. Tal Katz-Ezov at the Technion Genome Center, as well as Stern lab members for their support during an ongoing pandemic and various stages of lockdown. This work was funded by the Israeli Science Foundation (1333/16) and by a generous donation from the Milner foundation and from AppFlyer. This study was supported in part by a fellowship to DM, TK and OT from the Edmond J. Safra Center for Bioinformatics at Tel-Aviv University.

---

**Table S1.** Details of samples sequenced.

| Sample ID | Ct E | Ct RdRp | Ct N | Ct average | Sex | Age | Date | Hospital | Genome coverage | Geog. location of sample |
| --- | --- | --- | --- | --- | --- | --- | --- | --- | --- | --- |
| 1639953 | 28.5 | 30.5 | 32 | 30.18 | M | 10-19 | 26/3/20 | Poria | 0.996 | North |
| 1639996 | 23.1 | 24.4 | 26 | 24.45 | M | 20-29 | 26/3/20 | Poria | 0.996 | North |
| 2020038 | 22.5 |  |  | 22.46 | F | 50-59 | 17/3/20 | Hadassah | 0.994 | Tel Aviv |
| 2020051 | 30.9 |  |  | 30.87 | M | 40-49 | 17/3/20 | Hadassah | 0.976 | Tel Aviv |
| 2020063 | 28.4 |  |  | 28.39 | F | 20-29 | 17/3/20 | Hadassah | 0.985 | Jerusalem |
| 2020068 | 24.8 |  |  | 24.78 | F | 20-29 | 17/3/20 | Hadassah | 0.986 | Jerusalem |
| 2020069 | 28.2 |  |  | 28.24 | F | 30-39 | 17/3/20 | Hadassah | 0.987 | Jerusalem |
| 2020078 | 21.5 |  |  | 21.5 | M | 50-59 | 17/3/20 | Hadassah | 0.989 | Jerusalem |
| 2020084 | 24.9 |  |  | 24.87 | M | 60-69 | 17/3/20 | Hadassah | 0.994 | Jerusalem |
| 2020087 | 21.5 |  |  | 21.48 | M | 20-29 | 17/3/20 | Hadassah | 0.996 | Jerusalem |
| 2023920 | 18.6 |  |  | 18.56 | M | 40-49 | 17/3/20 | Hadassah | 0.996 | Jerusalem |
| 2023922 | 28 |  |  | 28 | F | 20-29 | 17/3/20 | Hadassah | 0.979 | Jerusalem |
| 2046129 | 23.1 | 24.2 | 27 | 24.88 | M | 20-29 | 31/3/20 | Poria | 0.996 | North |
| 2046171 | 29.6 | 30.8 | 33 | 31.24 | F | 50-59 | 31/3/20 | Poria | 0.954 | North |
| 2046434 | 19.5 | 20.6 | 24 | 21.28 | F | 50-59 | 1/4/20 | Poria | 0.996 | North |
| 2046548 | 28.5 | 31.1 | 34 | 31.01 | M | 40-49 | 1/4/20 | Poria | 0.991 | North |
| 2046614 | 24.4 | 26.2 | 28 | 26.24 | F | 30-39 | 2/4/20 | Poria | 0.996 | North |
| 2046616 | 20.5 | 22.3 | 25 | 22.47 | F | 70-79 | 2/4/20 | Poria | 0.996 | North |
| 2046815 | 13.8 | 16.5 | 18 | 16.08 | F | 80-89 | 4/4/20 | Poria | 0.994 | North |
| 2047004 | 23 | 24.5 | 27 | 24.81 | M | 30-39 | 27/3/20 | Poria | 0.996 | North |
| 2047011 | 28.5 | 30.2 | 32 | 30.24 | M | 20-29 | 26/3/20 | Poria | 0.952 | North |
| 2047016 | 25.7 | 26.7 | 29 | 26.99 | F | 0-9 | 26/3/20 | Poria | 0.996 | North |
| 2047145 | 10.3 | 11.8 | 15 | 12.38 | F | 30-39 | 26/3/20 | Poria | 0.996 | North |
| 2047188 | 13.6 | 15.3 | 18 | 15.48 | M | 20-29 | 26/3/20 | Poria | 0.996 | North |
| 2047189 | 9.75 | 11.7 | 14 | 11.87 | M | 20-29 | 26/3/20 | Poria | 0.996 | North |
| 2047364 | 20.8 | 22.8 | 25 | 22.68 | F | 60-69 | 28/3/20 | Poria | 0.996 | North |
| 2047392 | 27.6 | 29.9 | 31 | 29.42 | F | 30-39 | 29/3/20 | Poria | 0.966 | North |
| 2047418 | 24 | 25.4 | 28 | 25.67 | M | 0-9 | 29/3/20 | Poria | 0.996 | North |
| 2047567 | 26.8 | 28.7 | 31 | 28.97 | F | 30-39 | 29/3/20 | Poria | 0.978 | North |
| 2047586 | 22.8 | 24.1 | 27 | 24.71 | F | 20-29 | 29/3/20 | Poria | 0.996 | North |
| 2047604 | 22.7 | 24 | 26 | 24.32 | M | 50-59 | 29/3/20 | Poria | 0.996 | North |
| 2047738 | 20 | 22.1 | 25 | 22.21 | M | 10-19 | 30/3/20 | Poria | 0.996 | North |
| 2047749 | 19.3 | 20.7 | 23 | 21.03 | F | 40-49 | 30/3/20 | Poria | 0.996 | North |
| 2047772 | 23.8 | 25.7 | 26 | 25.3 | F | 30-39 | 30/3/20 | Poria | 0.996 | North |
| 2047883 | 21.1 | 22.9 | 25 | 22.87 | M | 60-69 | 31/3/20 | Poria | 0.996 | North |
| 2047927 | 32 | 33.7 | 36 | 33.74 | F | 60-69 | 31/3/20 | Poria | 0.918 | North |
| 2086001 | 28.4 |  |  | 28.4 | F | 10-19 | 1/4/20 | Hadassah | 0.988 | Jerusalem |
| 2086004 | 24.6 |  |  | 24.63 | F | 20-29 | 1/4/20 | Hadassah | 0.993 | Jerusalem |
| 2086008 | 20.2 |  |  | 20.24 | M | 90+ | 1/4/20 | Hadassah | 0.996 | Jerusalem |
| 2086012 | 24.4 |  |  | 24.39 | M | 20-29 | 1/4/20 | Hadassah | 0.988 | Jerusalem |
| 2086022 | 27.9 |  |  | 27.88 | M | 20-29 | 1/4/20 | Hadassah | 0.986 | Jerusalem |
| 2086033 | 28.8 |  |  | 28.79 | F | 60-69 | 1/4/20 | Hadassah | 0.991 | Jerusalem |
| 2086034 | 30 |  |  | 30.01 | F | 20-29 | 1/4/20 | Hadassah | 0.981 | Jerusalem |
| 2086045 | 20 |  |  | 19.99 | M | 50-59 | 1/4/20 | Hadassah | 0.996 | Jerusalem |
| 2089366 | 20.4 |  |  | 20.36 | M | 20-29 | 1/4/20 | Hadassah | 0.996 | Jerusalem |
| 2089368 | 25.6 |  |  | 25.62 | M | 60-69 | 1/4/20 | Hadassah | 0.985 | Jerusalem |
| 2089375 | 27.5 |  |  | 27.54 | M | 10-19 | 1/4/20 | Hadassah | 0.992 | Jerusalem |
| 2089380 | 30.1 |  |  | 30.14 | F | 60-69 | 1/4/20 | Hadassah | 0.979 | Jerusalem |
| 2089383 | 29.8 |  |  | 29.79 | M | 30-39 | 1/4/20 | Hadassah | 0.989 | Jerusalem |
| 2089697 | 28.3 |  |  | 28.27 | F | 20-29 | 1/4/20 | Hadassah | 0.989 | Jerusalem |
| 2089698 | 28.6 |  |  | 28.58 | F | 50-59 | 1/4/20 | Hadassah | 0.984 | Jerusalem |
| 2089712 | 25.5 |  |  | 25.52 | M | 20-29 | 1/4/20 | Hadassah | 0.989 | Jerusalem |
| 2089718 | 20.6 |  |  | 20.6 | F | 30-39 | 1/4/20 | Hadassah | 0.996 | Jerusalem |
| 2089723 | 28.6 |  |  | 28.55 | M | 30-39 | 1/4/20 | Hadassah | 0.992 | Jerusalem |
| 2089812 | 23.3 |  |  | 23.29 | F | 20-29 | 1/4/20 | Hadassah | 0.995 | Jerusalem |
| 2089839 | 25.5 |  |  | 25.5 | M | 60-69 | 1/4/20 | Hadassah | 0.989 | Jerusalem |
| 2089852 | 25.5 |  |  | 25.53 | F | 60-69 | 1/4/20 | Hadassah | 0.989 | Jerusalem |
| 2089861 | 18.3 |  |  | 18.31 | M | 70-79 | 1/4/20 | Hadassah | 0.996 | Jerusalem |
| 2089863 | 18.8 |  |  | 18.8 | M | 30-39 | 1/4/20 | Hadassah | 0.996 | Jerusalem |
| 2089866 | 20.7 |  |  | 20.7 | M | 30-39 | 1/4/20 | Hadassah | 0.996 | Jerusalem |
| 2099018 | 30.3 |  |  | 30.34 | F | 60-69 | 2/4/20 | Hadassah | 0.980 | Jerusalem |
| 2099019 | 27.5 |  |  | 27.48 | M | 20-29 | 2/4/20 | Hadassah | 0.992 | Jerusalem |
| 2099159 | 27.3 |  |  | 27.3 | F | 10-19 | 2/4/20 | Hadassah | 0.988 | Jerusalem |
| 2099251 | 23 |  |  | 22.98 | F | 20-29 | 2/4/20 | Hadassah | 0.996 | Jerusalem |
| 2099416 | 29 |  |  | 28.98 | M | 20-29 | 2/4/20 | Hadassah | 0.983 | Jerusalem |
| 2099421 | 20.5 |  |  | 20.45 | M | 50-59 | 2/4/20 | Hadassah | 0.996 | Jerusalem |
| 2107132 | 29.9 |  |  | 29.91 | F | 50-59 | 14/4/20 | Hadassah | 0.996 | Jerusalem |
| 2107137 | 19.6 |  |  | 19.6 | F | 10-19 | 14/4/20 | Hadassah | 0.996 | Jerusalem |
| 2107681 | 22.3 |  |  | 22.3 | F | 30-39 | 14/4/20 | Hadassah | 0.996 | Jerusalem |
| 2113155 | 25.6 |  |  | 25.55 | M | 0-9 | 15/4/20 | Hadassah | 0.996 | Jerusalem |
| 2113161 | 25.9 |  |  | 25.92 | M | 70-79 | 15/4/20 | Hadassah | 0.996 | Jerusalem |
| 2113173 | 29.8 |  |  | 29.78 | M | 10-19 | 15/4/20 | Hadassah | 0.996 | Jerusalem |
| 2113174 | 26.4 |  |  | 26.42 | M | 70-79 | 15/4/20 | Hadassah | 0.996 | Tel Aviv |
| 2113178 | 26.6 |  |  | 26.62 | M | 20-29 | 15/4/20 | Hadassah | 0.996 | Jerusalem |
| 2113255 | 29.1 |  |  | 29.14 | F | 60-69 | 14/4/20 | Hadassah | 0.994 | Jerusalem |
| 2113256 | 29.7 |  |  | 29.65 | F | 10-19 | 14/4/20 | Hadassah | 0.996 | Jerusalem |
| 2113601 | 25.1 |  |  | 25.07 | M | 0-9 | 15/4/20 | Hadassah | 0.996 | Jerusalem |
| 2113603 | 21.7 |  |  | 21.72 | M | 70-79 | 15/4/20 | Hadassah | 0.996 | Jerusalem |
| 2113678 | 29.8 |  |  | 29.78 | M | 0-9 | 15/4/20 | Hadassah | 0.996 | Tel Aviv |
| 2115701 | 26 |  |  | 25.96 | F | 10-19 | 16/4/20 | Hadassah | 0.996 | Jerusalem |
| 2115964 | 28.4 |  |  | 28.35 | M | 10-19 | 16/4/20 | Hadassah | 0.996 | Tel Aviv |
| 2115968 | 26.8 |  |  | 26.83 | F | 20-29 | 16/4/20 | Hadassah | 0.996 | Tel Aviv |
| 2115976 | 28.2 |  |  | 28.17 | M | 60-69 | 16/4/20 | Hadassah | 0.985 | Tel Aviv |
| 2115980 | 24.5 |  |  | 24.49 | F | 20-29 | 16/4/20 | Hadassah | 0.912 | North |
| 2115990 | 30.2 |  |  | 30.15 | F | 50-59 | 16/4/20 | Hadassah | 0.996 | Jerusalem |
| 2116859 | 29.6 |  |  | 29.6 | F | 10-19 | 20/4/20 | Hadassah | 0.953 | Jerusalem |
| 2123853 | 28.1 |  |  | 28.05 | M | 70-79 | 20/4/20 | Hadassah | 0.996 | Jerusalem |
| 2123863 | 27.5 |  |  | 27.47 | M | 80-89 | 20/4/20 | Hadassah | 0.996 | Jerusalem |
| 13075703 | 24 | 25.6 | 28 | 25.98 | F | 20-29 | 29/3/20 | Sheba | 0.996 | Tel Aviv |
| 13075719 | 27.7 | 28.9 | 31 | 29.16 | F | 20-29 | 29/3/20 | Sheba | 0.996 | Tel Aviv |
| 13075735 | 26.8 | 28.4 | 30 | 28.47 | M | 40-49 | 30/3/20 | Sheba | 0.996 | Tel Aviv |
| 13075782 | 29.1 | 30.5 | 33 | 30.85 | F | 40-49 | 30/3/20 | Sheba | 0.992 | Tel Aviv |
| 13075788 | 27.8 | 29.3 | 31 | 29.4 | F | 40-49 | 30/3/20 | Sheba | 0.996 | Tel Aviv |
| 13075790 | 27.2 | 28.8 | 30 | 28.7 | M | 50-59 | 30/3/20 | Sheba | 0.996 | Tel Aviv |
| 13075832 | 17.5 | 18.6 | 20 | 18.7 | M | 30-39 | 30/3/20 | Sheba | 0.996 | Tel Aviv |
| 13075879 | 18.6 | 20 | 22 | 20.27 | M | 50-59 | 30/3/20 | Sheba | 0.993 | Tel Aviv |
| 13075882 | 18.6 | 20.4 | 21 | 20.1 | F | 20-29 | 30/3/20 | Sheba | 0.996 | Tel Aviv |
| 13075914 | 17.1 | 18.5 | 20 | 18.39 | F | 50-59 | 30/3/20 | Sheba | 0.996 | Tel Aviv |
| 13077377 | 27 | 29 | 29 | 28.33 | M | 40-49 | 2/4/20 | Sheba | 0.996 | Tel Aviv |
| 13077383 | 26 | 26 | 28 | 26.67 | F | 40-49 | 2/4/20 | Sheba | 0.984 | Jerusalem |
| 13077413 | 24 | 25 | 26 | 25 | M | 70-79 | 3/4/20 | Sheba | 0.996 | Tel Aviv |
| 13077494 | 28 | 29 | 31 | 29.33 | M | 20-29 | 2/4/20 | Sheba | 0.856 | South Coast |
| 13077497 | 21 | 22 | 24 | 22.33 | F | 60-69 | 2/4/20 | Sheba | 0.996 | South Coast |
| 13077498 | 17 | 18 | 21 | 18.67 | M | 20-29 | 3/4/20 | Sheba | 0.996 | South Coast |
| 13077510 | 21 | 22 | 25 | 22.67 | M | 50-59 | 3/4/20 | Sheba | 0.993 | Jerusalem |
| 13077511 | 25 | 26 | 28 | 26.33 | M | 20-29 | 3/4/20 | Sheba | 0.986 | Jerusalem |
| 13077558 | 19 | 20 | 22 | 20.33 | F | 30-39 | 3/4/20 | Sheba | 0.996 | Tel Aviv |
| 13077560 | 27 | 28 | 30 | 28.33 | F | 80-89 | 3/4/20 | Sheba | 0.974 | Tel Aviv |
| 13077562 | 15 | 17 | 19 | 17 | F | 50-59 | 3/4/20 | Sheba | 0.988 | Tel Aviv |
| 13077564 | 22 | 23 | 25 | 23.33 | F | 50-59 | 3/4/20 | Sheba | 0.992 | Tel Aviv |
| 13077711 | 27 | 28 | 30 | 28.33 | F | 30-39 | 3/4/20 | Sheba | 0.995 | Tel Aviv |
| 13077723 | 22 | 23 | 25 | 23.33 | M | ? | 3/4/20 | Sheba | 0.989 | Tel Aviv |
| 13077726 | 20 | 20 | 22 | 20.67 | F | 60-69 | 3/4/20 | Sheba | 0.996 | Tel Aviv |
| 13077803 | 28 | 29 | 32 | 29.67 | F | 30-39 | 4/4/20 | Sheba | 0.981 | South Coast |
| 13077823 | 15 | 17 | 19 | 17 | F | 10-19 | 4/4/20 | Sheba | 0.996 | South Coast |
| 13077840 | 23 | 25 | 27 | 25 | M | 20-29 | 4/4/20 | Sheba | 0.991 | Jerusalem |
| 13077846 | 19 | 20 | 23 | 20.67 | F | 0-9 | 4/4/20 | Sheba | 0.996 | Tel Aviv |
| 13077847 | 24 | 26 | 27 | 25.67 | M | 10-19 | 4/4/20 | Sheba | 0.986 | Tel Aviv |
| 13077875 | 18 | 19 | 21 | 19.33 | F | 40-49 | 4/4/20 | Sheba | 0.996 | Tel Aviv |
| 13077882 | 24 | 26 | 27 | 25.67 | M | 40-49 | 4/4/20 | Sheba | 0.987 | Jerusalem |
| 51137031 |  |  |  | 18.58 | M | 30-39 | 1/3/20 | Soroka | 0.996 | Tel Aviv |
| 51137844 |  |  |  | 27.6 | F | 30-39 | 17/3/20 | Soroka | 0.983 | South |
| 51140028 |  |  |  | 20.56 | F | 50-59 | 17/3/20 | Soroka | 0.996 | North |
| 51140068 |  |  |  | 22.8 | M | 30-39 | 18/3/20 | Soroka | 0.988 | South |
| 51140271 |  |  |  | 28.16 | F | 40-49 | 18/3/20 | Soroka | 0.971 | South Coast |
| 51140279 |  |  |  | 25.76 | F | 20-29 | 18/3/20 | Soroka | 0.976 | South Coast |
| 51140315 |  |  |  | 23.41 | M | 60-69 | 18/3/20 | Soroka | 0.996 | South |
| 51140539 |  |  |  | 19.4 | F | 50-59 | 19/3/20 | Soroka | 0.996 | South |
| 51140836 |  |  |  | 24.5 | M | 30-39 | 20/3/20 | Soroka | 0.986 | South |
| 51141014 |  |  |  | 23.08 | F | 90+ | 22/3/20 | Soroka | 0.996 | South |
| 51141121 |  |  |  | 29.05 | M | 70-79 | 22/3/20 | Soroka | 0.989 | South |
| 51141225 |  |  |  | 18.58 | F | 40-49 | 22/3/20 | Soroka | 0.996 | South |
| 51144342 |  |  |  | 25.59 | M | 90+ | 30/3/20 | Soroka | 0.976 | South |
| 51145198 |  |  |  | 25.49 | M | 80-89 | 1/4/20 | Soroka | 0.989 | South |
| 51145482 |  |  |  | 27.92 | F | 40-49 | 5/4/20 | Soroka | 0.992 | Tel Aviv |
| 51146355 |  |  |  | 29.24 | F | 80-89 | 4/4/20 | Soroka | 0.989 | South |
| 51146500 |  |  |  | 21.79 | M | 40-49 | 5/4/20 | Soroka | 0.989 | Tel Aviv |
| 51146503 |  |  |  | 23.75 | F | 30-39 | 5/4/20 | Soroka | 0.993 | Jerusalem |
| 51146669 |  |  |  | 24.41 | M | 10-19 | 5/4/20 | Soroka | 0.994 | Tel Aviv |
| 51146683 |  |  |  | 24.8 | M | 10-19 | 5/4/20 | Soroka | 0.965 | Tel Aviv |
| 130710062 | 28 | 28 | 28 | 28 | M | 40-49 | 14/4/20 | Sheba | 0.991 | Tel Aviv |
| 130710067 | 23 | 24 | 24 | 23.67 | M | 60-69 | 14/4/20 | Sheba | 0.996 | Tel Aviv |
| 130710097 | 29 | 31 | 32 | 30.67 | M | 70-79 | 15/4/20 | Sheba | 0.996 | Tel Aviv |
| 130710099 | 33 | 33 | 34 | 33.33 | M | 60-69 | 15/4/20 | Sheba | 0.979 | Tel Aviv |
| 130710157 | 20 | 21 | 24 | 21.67 | M | 70-79 | 15/4/20 | Sheba | 0.996 | Tel Aviv |
| 130710159 | 27 | 28 | 29 | 28 | F | 80-89 | 15/4/20 | Sheba | 0.990 | Tel Aviv |
| 130710211 | 18 | 19 | 20 | 19 | F | 70-79 | 16/4/20 | Sheba | 0.996 | Tel Aviv |
| 130710217 | 22 | 23 | 26 | 23.67 | M | 50-59 | 16/4/20 | Sheba | 0.996 | Tel Aviv |
| 130710390 | 28 | 29 | 30 | 29 | F | 30-39 | 17/4/20 | Sheba | 0.996 | Tel Aviv |
| 130710414 | 20 | 21 | 22 | 21 | M | 20-29 | 17/4/20 | Sheba | 0.996 | Tel Aviv |
| 130710643 | 31 | 31 | 31 | 31 | M | 40-49 | 19/4/20 | Sheba | 0.986 | Tel Aviv |
| 130710644 | 29 | 30 | 31 | 30 | F | 80-89 | 19/4/20 | Sheba | 0.985 | Tel Aviv |
| 130710716 | 30 | 31 | 31 | 30.67 | F | 60-69 | 19/4/20 | Sheba | 0.989 | Tel Aviv |
| 130711082 | 18.5 | 20 | 22 | 20.17 | F | 60-69 | 20/4/20 | Sheba | 0.996 | Tel Aviv |
| 130711104 | 27 | 29 | 29 | 28.33 | F | 80-89 | 20/4/20 | Sheba | 0.993 | Tel Aviv |
| 130711112 | 31 | 32 | 31 | 31.33 | F | 80-89 | 21/4/20 | Sheba | 0.991 | Tel Aviv |
| 130711116 | 25 | 27 | 25 | 25.67 | F | 60-69 | 21/4/20 | Sheba | 0.996 | Tel Aviv |
| 130711367 | 29 | 27 | 27 | 27.67 | F | 60-69 | 22/4/20 | Sheba | 0.996 | North |
| 130711417 | 32 | 31 | 30 | 31 | M | 60-69 | 22/4/20 | Sheba | 0.974 | Tel Aviv |
| 701002313 |  |  |  | 27 | M | 50-59 | 21/3/20 | Barzilai | 0.996 | South Coast |
| 701002314 |  |  |  | 21 | M | 20-29 | 21/3/20 | Barzilai | 0.996 | South Coast |
| 701002317 |  |  |  | 25 | M | 50-59 | 21/3/20 | Barzilai | 0.996 | South Coast |
| 701002327 |  |  |  | 19 | M | 50-59 | 22/3/20 | Barzilai | 0.996 | South Coast |
| 701002334 |  |  |  | 21 | M | 70-79 | 22/3/20 | Barzilai | 0.996 | South Coast |
| 701002403 |  |  |  | 31 | M | 40-49 | 23/3/20 | Barzilai | 0.996 | South Coast |
| 701002407 |  |  |  | 27 | M | 40-49 | 23/3/20 | Barzilai | 0.959 | South Coast |
| 701002426 |  |  |  | 25 | M | 60-69 | 24/3/20 | Barzilai | 0.996 | South Coast |
| 701002431 |  |  |  | 18 | M | 20-29 | 24/3/20 | Barzilai | 0.996 | South Coast |
| 701002440 |  |  |  | 26 | F | 20-29 | 24/3/20 | Barzilai | 0.996 | South Coast |
| 701002442 |  |  |  | 18 | M | 30-39 | 24/3/20 | Barzilai | 0.996 | South Coast |
| 701002455 |  |  |  | 30 | F | 20-29 | 24/3/20 | Barzilai | 0.992 | South Coast |
| 701002456 |  |  |  | 30 | F | 60-69 | 24/3/20 | Barzilai | 0.980 | South Coast |
| 701002458 |  |  |  | 21 | M | 40-49 | 24/3/20 | Barzilai | 0.996 | South Coast |
| 701002462 |  |  |  | 21 | M | 70-79 | 24/3/20 | Barzilai | 0.996 | South Coast |
| 701002489 |  |  |  | 27 | M | 40-49 | 25/3/20 | Barzilai | 0.996 | South Coast |
| 701002504 |  |  |  | 21 | M | 70-79 | 25/3/20 | Barzilai | 0.996 | South Coast |
| 701002538 |  |  |  | 28 | M | 70-79 | 25/3/20 | Barzilai | 0.996 | South Coast |
| 701002540 |  |  |  | 21 | F | 40-49 | 25/3/20 | Barzilai | 0.996 | South Coast |
| 701002550 |  |  |  | 24 | M | 30-39 | 26/3/20 | Barzilai | 0.996 | South Coast |
| 701002555 |  |  |  | 23 | F | 40-49 | 26/3/20 | Barzilai | 0.996 | South Coast |
| 701002556 |  |  |  | 22 | F | 60-69 | 26/3/20 | Barzilai | 0.996 | South Coast |
| 701002561 |  |  |  | 28 | M | 50-59 | 26/3/20 | Barzilai | 0.996 | South Coast |
| 701002591 |  |  |  | 24 | F | 50-59 | 26/3/20 | Barzilai | 0.996 | South Coast |
| 701002666 |  |  |  | 26 | M | 40-49 | 26/3/20 | Barzilai | 0.996 | South Coast |
| 701002681 |  |  |  | 24 | M | 60-69 | 27/3/20 | Barzilai | 0.996 | South Coast |
| 701002752 |  |  |  | 27 | M | 10-19 | 28/3/20 | Barzilai | 0.987 | South Coast |
| 701002768 |  |  |  | 21 | M | 50-59 | 28/3/20 | Barzilai | 0.996 | South Coast |
| 701002786 |  |  |  | 27 | M | 60-69 | 29/3/20 | Barzilai | 0.996 | South Coast |
| 701002792 |  |  |  | 25 | M | 50-59 | 29/3/20 | Barzilai | 0.995 | South Coast |
| 990059202 | 26.9 | 28.4 | 27 | 27.4 | M | 60-69 | 25/3/20 | Assuta | 0.996 | South Coast |
| 990059203 | 26.5 | 28.6 | 27 | 27.2 | F | 80-89 | 25/3/20 | Assuta | 0.996 | South Coast |
| 990059204 | 28.4 | 30.1 | 28 | 28.97 | F | 30-39 | 25/3/20 | Assuta | 0.976 | South Coast |
| 990059217 | 29.1 | 31.4 | 32 | 30.9 | M | 60-69 | 25/3/20 | Assuta | 0.995 | South Coast |
| 990059230 | 26.5 | 29.7 | 29 | 28.5 | F | 0-9 | 25/3/20 | Assuta | 0.972 | South Coast |
| 990059231 | 24.9 | 28 | 28 | 27 | M | 30-39 | 25/3/20 | Assuta | 0.996 | South Coast |
| 990059232 | 24.5 | 26.7 | 28 | 26.3 | M | 10-19 | 25/3/20 | Assuta | 0.996 | South Coast |
| 990059233 | 19.7 | 21.8 | 23 | 21.43 | F | 50-59 | 25/3/20 | Assuta | 0.996 | South Coast |
| 990059237 | 28.3 | 30.6 | 32 | 30.23 | F | 0-9 | 25/3/20 | Assuta | 0.989 | South Coast |
| 990059238 | 24.3 | 26.3 | 27 | 25.83 | F | 20-29 | 25/3/20 | Assuta | 0.996 | South Coast |
| 990059244 | 27.9 | 30.8 | 31 | 29.97 | F | ? | 25/3/20 | Assuta | 0.938 | South Coast |
| 990059251 | 24.7 | 28 | 28 | 26.9 | F | 50-59 | 25/3/20 | Assuta | 0.996 | South Coast |
| 990059252 | 24.4 | 26.8 | 28 | 26.27 | F | 20-29 | 25/3/20 | Assuta | 0.996 | South Coast |
| 990300681 | 22.9 | 26.4 | 27 | 25.37 | F | 60-69 | 30/3/20 | Assuta | 0.995 | South Coast |
| 990300691 | 29.3 | 31.7 | 32 | 31 | M | 20-29 | 31/3/20 | Assuta | 0.970 | South Coast |
| 990300724 | 23.7 | 26.6 | 26 | 25.53 | M | 80-89 | 1/4/20 | Assuta | 0.993 | South Coast |
| 990300860 | 19.3 | 21.9 | 0 | 20.6 | M | 20-29 | 2/4/20 | Assuta | 0.996 | South Coast |
| 990307712 | 26 | 27.3 | 28 | 26.97 | F | 20-29 | 26/3/20 | Assuta | 0.996 | South Coast |
| 990333068 | 30.1 | 32.7 | 34 | 32.13 | M | 60-69 | 24/3/20 | Assuta | 0.963 | South Coast |
| 990333189 | 29.3 | 31.4 | 32 | 30.9 | M | 50-59 | 26/3/20 | Assuta | 0.950 | South Coast |
| 990333193 | 20.7 | 22.6 | 24 | 22.43 | F | 40-49 | 26/3/20 | Assuta | 0.996 | South Coast |
| 990333263 | 36.4 | 0 | 0 | 36.4 | M | 30-39 | 31/3/20 | Assuta | 0.660 | South Coast |
| 990430264 | 29.3 | 31.7 | 33 | 31.17 | F | 40-49 | 29/3/20 | Assuta | 0.970 | South Coast |
| 990430265 | 29.6 | 32.2 | 33 | 31.7 | F | 30-39 | 29/3/20 | Assuta | 0.994 | South Coast |

**Table S2.** Model parameter priors and estimated values.

| Parameter | Prior | Posterior (Median (95% HPD)) |  |  |  |  |  |  |
| --- | --- | --- | --- | --- | --- | --- | --- | --- |
| | | $p_h = 0.01$ | $p_h = 0.02$ | $p_h = 0.05$ | $p_h = 0.07$ | $p_h = 0.08$ | $p_h = 0.09$ | $p_h = 0.10$ |
| Clock rate | Uniform(0.0007, 0.002) | 7.7E-4<br>(7.0E-4,<br>8.4E-4) | 7.3E-4 (7.0E-4,<br>8.1E-4) | 7.4E-4 (7.0E-4,<br>8.2E-4) | 7.5E-4 (7.0E-4,<br>8.5E-4) | 7.8E-4 (7.1E-4,<br>8.7E-4) | 7.5E-4 (7.0E-4,<br>8.6E-4) | 7.5E-4 (7.0E-4,<br>8.2E-4) |
| $\kappa$ | Lognormal(mean = 1.0, sd = 1.25) | 3.7 (3.1, 4.5) | 3.7 (3.0, 4.4) | 3.7 (3.0, 4.4) | 3.7 (3.0, 4.5) | 3.7 (3.1, 4.5) | 3.7 (3.1, 4.5) | 3.7 (3.0, 4.3) |
| $\gamma$ | Exponential(mean = 1.0) | 0.039 (0.003, 0.082) | 0.047 (0.003, 0.109) | 0.040 (0.002, 0.104) | 0.042 (0.003, 0.010) | 0.039 (0.002, 0.098) | 0.039 (0.005, 0.088) | 0.034 (0.003, 0.080) |
| $R_0$ | Lognormal(mean = 1.5, sd = 0.5) | 2.7 (2.5, 2.9) | 2.4 (2.3, 2.5) | 2.1 (2.0, 2.3) | 2.1 (1.9, 2.2) | 2.0 (1.9, 2.2) | 2.0 (1.9, 2.2) | 2.1 (1.9, 2.2) |
| $\alpha$ | Uniform(0, 2) | 0.26 (0.15, 0.49) | 0.30 (0.15, 0.51) | 0.27 (0.15, 0.49) | 0.36 (0.19, 0.63) | 0.36 (0.16, 0.66) | 0.32 (0.18, 0.58) | 0.32 (0.16, 0.65) |
| $E_{init}$ | Exponential(mean = 1.0) | 21 (14, 30) | 23 (19, 27) | 21 (15, 32) | 19 (14, 26) | 18 (12, 27) | 17 (12, 23) | 12 (9, 16) |
| $Y_{init}$ | Exponential(mean = 1.0) | 0.021 (0.018, 0.036) | 0.029 (0.029, 0.049) | 0.080 (0.026, 0.122) | 0.018 (0.012, 0.035) | 0.015 (0.012, 0.019) | 0.056 (0.040, 0.078) | 0.12 (0.07, 0.19) |
| $\rho$ | Lognormal(mean = 3.6, sd = 1.0) | 27 (25, 28) | 25 (24, 26) | 21 (20, 24) | 23 (22, 25) | 23 (22, 24) | 21 (20, 22) | 19 (18, 20) |
| $\eta$ | Exponential(mean = 10 ) | 9.4 (6.7, 10.0) | 9.7 (8.3, 10.0) | 9.5 (7.4, 10.0) | 9.5 (7.5, 10.0) | 9.2 (6.8, 10.0) | 9.5 (7.6, 10.0) | 9.7 (8.1, 10.0) |

